# Ebola Disease: Knowledge, attitude, and practices among medical students at a tertiary institution in Uganda

**DOI:** 10.1101/2024.11.05.24316752

**Authors:** Daniel Gakwerere, Emmanuel Biryabarema, Melanie Namubiru, Marjorine Namuyomba, Bridget Atuhaire, Carol Musubika, David Mukunya

**Affiliations:** School of Medicine, College of Health Sciences, Makerere University, Kampala, Uganda; Department of Immunology and Molecular Biology, Makerere University, Kampala, Uganda; Department of Community and Public Health, Busitema University, Mbale, Uganda

## Abstract

**Background:** Ebolavirus Disease (EVD) has been a public health threat since its discovery in 1976 with occasional outbreaks on the African continent. This study sought to assess the knowledge, attitudes and practices (KAP) among medical students at a tertiary university in Uganda.

**Methods:** We conducted a descriptive cross-sectional study using WhatsApp Messenger among students in their clinical years. A pre-validated questionnaire was adapted and modified to assess KAP towards EVD. All analyses were performed using Statistical Package for Social Sciences (SPSS) version 29.0. Bloom’s cutoff of 80% was used to assess sufficient knowledge (≥80%), good attitude (≥6.4) and good practices (≥4.8).

**Results:** A total of 339 participated, majority were males (n=210, 61.9%) with mean age of 24.2 years (Standard Deviation: 4.0). 77.9% (n=264) were pursuing Bachelor of Medicine and Bachelor of Surgery. Overall, 13.6% (n=46) had sufficient knowledge, 10.9% (n=37) had good attitude and 32.7% (n=111) had good practices. 54.9% (n=186) always avoided patients with signs and symptoms suggestive of EVD while 11.8% (n=40) would accept an approved ebolavirus vaccine.

**Conclusion:** These results revealed suboptimal EVD-related KAP among medical students. We recommend training of students on clinical presentation, transmission, treatment and prevention of EVD to effectively control future outbreaks.

## Introduction

Ebola Virus Disease (EVD) is a rare and deadly haemorrhagic disease in people and nonhuman primates (1). Filovirus belonging to the family Filoviridae cause EVD. Six different species of ebolaviruses have been identified: *Zaire ebolavirus* (EBOV), *Sudan ebolavirus* (SUDV), *Bundibugyo ebolavirus* (BDBV), *Tai Forest ebolavirus* (TAFV) *Reston ebolavirus* (RESTV) and the newly discovered *Bombali ebolavirus* (BOMV) (2,3).

Contact with infected animals such as Fruit bats and monkeys is thought to be the initial source of infection. The virus spreads from person to person through direct contact with blood and other body fluids. The disease manifests with fever, malaise, sore throat, gastrointestinal symptoms and unexplained bleeding (4). This deadly disease has recorded occasional outbreaks occurring mostly on the African continent since the first cases were detected in 1976 when two concomitant outbreaks happened in Sudan and Democratic Republic of Congo. Sudan ebolavirus caused the fifth and most recent outbreak of EVD in Uganda (5). It is also responsible for the largest outbreak that resulted in 425 cases and 224 deaths (6).

Subsequently, there have been several outbreaks in many countries mostly in Africa, i.e. Democratic Republic of Congo, Gabon, Guinea, Liberia, Sierra Leone, Sudan and Uganda. Imported cases have been reported in Mali, Nigeria, Senegal, Europe and United States of America (7).

Sporadic outbreaks have occurred in Uganda since 2000, recorded as the first and largest with 53% Case Fatality Rate (CFR) (6). The most recent outbreak between August 8 and November 27, 2022 where 164 cases of SUVD. 77 died (47% CFR) and 87 recovered (8).

During this outbreak, at least 19 Health Care Workers (HCWs) were infected, of whom 7 died (9). This outbreak followed a global pandemic of Covid 19 which strained Uganda’s health system, killing 37 HCWs (10).

Most studies on KAPs involving HCWs in health facilities across Africa have been focused on doctors, nurses, laboratory technicians and pharmacists. To our knowledge, there is paucity of data on KAP of medical students who are the future healthcare system and frontline workers towards EVD. The purpose of the study was to assess the Knowledge Attitude and Practices towards Ebola Virus Disease of medical students in their clinical years at Makerere university in Uganda toward EVD.

## Materials and Methods

### Study design and site

We conducted a cross-sectional study from December 2023 to March 2024 at Makerere University College of Health Sciences (MakCHS), Makerere University, Uganda, Kampala. The institution is one of the oldest and among the top medical schools in Africa. MakCHS has four schools offering 12 undergraduate programs of approximately 4,000 students as of Academic Year 2022/2023. The duration of courses offered ranges from three to five years.

### Study population

Medical students admitted to MakCHS in their clinical years offering; Bachelor of Medicine and Surgery (MBChB, 5 years), Bachelor of Dental Surgery (BDS, 5years) and Bachelor of Nursing (BNUR, 4 years) and had finished at least one clinical rotation were included in the study after an informed consent.

### Study procedure

Participants identified to represent the proportion of each year and course in the study were approached using WhatsApp Messenger (Facebook, Inc., California, USA) for their participation. A total of 391 students were approached to participate in the study. An online data collection tool was designed and executed using Kobotool (via kf.kobotoolbox.org). The link to the questionnaire was sent to the enrolled participants via their identified WhatsApp numbers. Written informed consent was sought for every participant before proceeding to fill the questionnaire. The study was conducted between December 19, 2023 and March 21, 2024.

### Operational definition

Medical students are individuals enrolled in a medical school at a university. Medical students in third year of medical school, start periodical rotations in internal medicine, surgery, paediatrics, obstetrics and gynaecology among others for their clinical clerkship (11). This training is mainly in teaching hospitals which double as referral facilities in Uganda (12) and rural health facilities for Community-Based Education and Research (COBERS) (13).

This group of people become healthcare workers at the end of their training. For the purpose of this study, medical students in their clinical years of study that interface with patients were enrolled. These included BNUR ( in their 3^rd^ and 4^th^ years), BDS ( in their 3^rd^, 4^th^ and 5^th^ years) MBChB ( in their 3^rd^, 4^th^ and 5^th^ years).

### Study variables

#### Independent variables

Demographic details such as age, sex, year of study, academic program, and ward rotation completed.

#### Dependent variables

Knowledge, attitude and practices toward EVD.

Knowledge was assessed using a 6-item questionnaire adapted from Jalloh *et al.* and modified to outfit our study. A weight of 1 point was attached to each correct answer. The questions were about source of information about EVD, transmission, and clinical presentations. Each correct response weight 1 point and 0 for incorrect responses. The higher the points, the more knowledgeable the student is.

Attitudes were assessed using 6 Likert-item questions that have been adopted from Olum *et al* and modified suitably for our study by the authors. The responses were; strongly disagree, disagree, neutral, agree and strongly agree each weighing 1–5 respectively for each positive statement.

Practices were assessed using 6 Likert-item questions that have been developed from the WHO recommended practices for prevention of EVD transmission i.e., washing hands, avoiding crowded places, avoiding handshakes and using PPEs. The responses were; always, occasional, and never each weighing 3, 2, and 1 point respectively for a good practice.

### Data management and analyses

Completed questionnaires were extracted from Kobotool and exported to a Microsoft Excel 2016 for cleaning and coding. The cleaned data was exported to SPSS version: 29.0 for analyses. Numerical data was summarized as means and standard deviations or median and range as appropriate. Categorical data was summarized as frequencies and proportions. Bloom’s cut-off of 80% was used to determine sufficient knowledge (≥80%), positive attitude(≥6.4), and good practice (≥4.8). Associations between independent variables and dependent variables were assessed using multivariate analysis in SPSS version: 29.0 software. A p < 0.05 is considered statistically significant.

## Results

Of the 391 participants approached, only 339 agreed to participate in the study. Majority of them were male (n= 210, 61.9%), with mean age of 24.2 (Standard Deviation: 4.0) years and mostly 21-30 (n= 310, 91.4%) years. A vast majority were pursuing MBChB (n= 264, 77.9%) and year 3 medical students participated more (n= 141, 41.6%). All the participants had completed at least one of the four major rotations in medicine. Information regarding EVD was mainly obtained from news media, social media like WhatsApp and Twitter, official government sites and media plus official international health sites and media like CDC and WHO. **Table 1** summarises the sociodemographic characteristics of the participants.

**TABLE 1.**
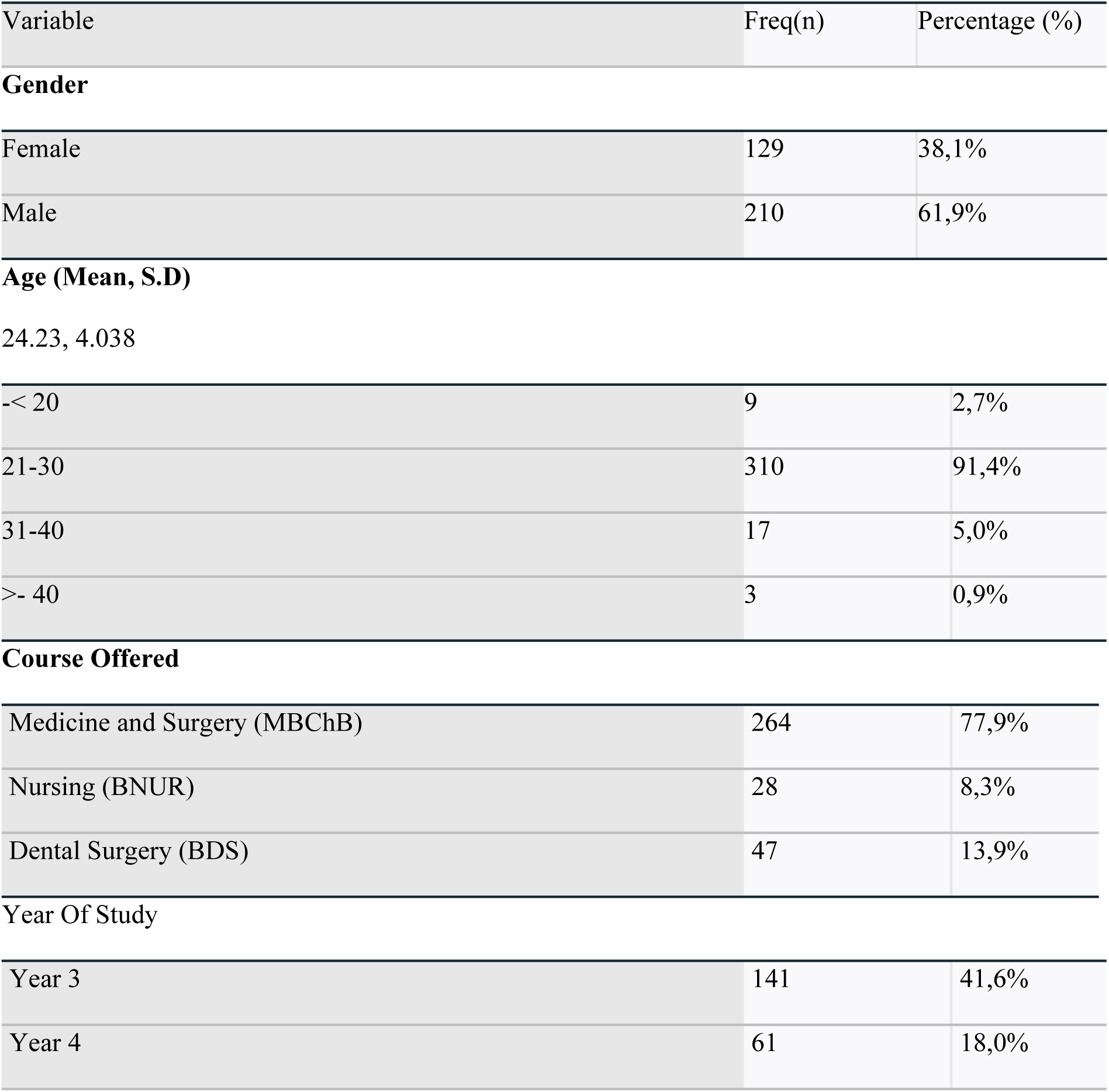
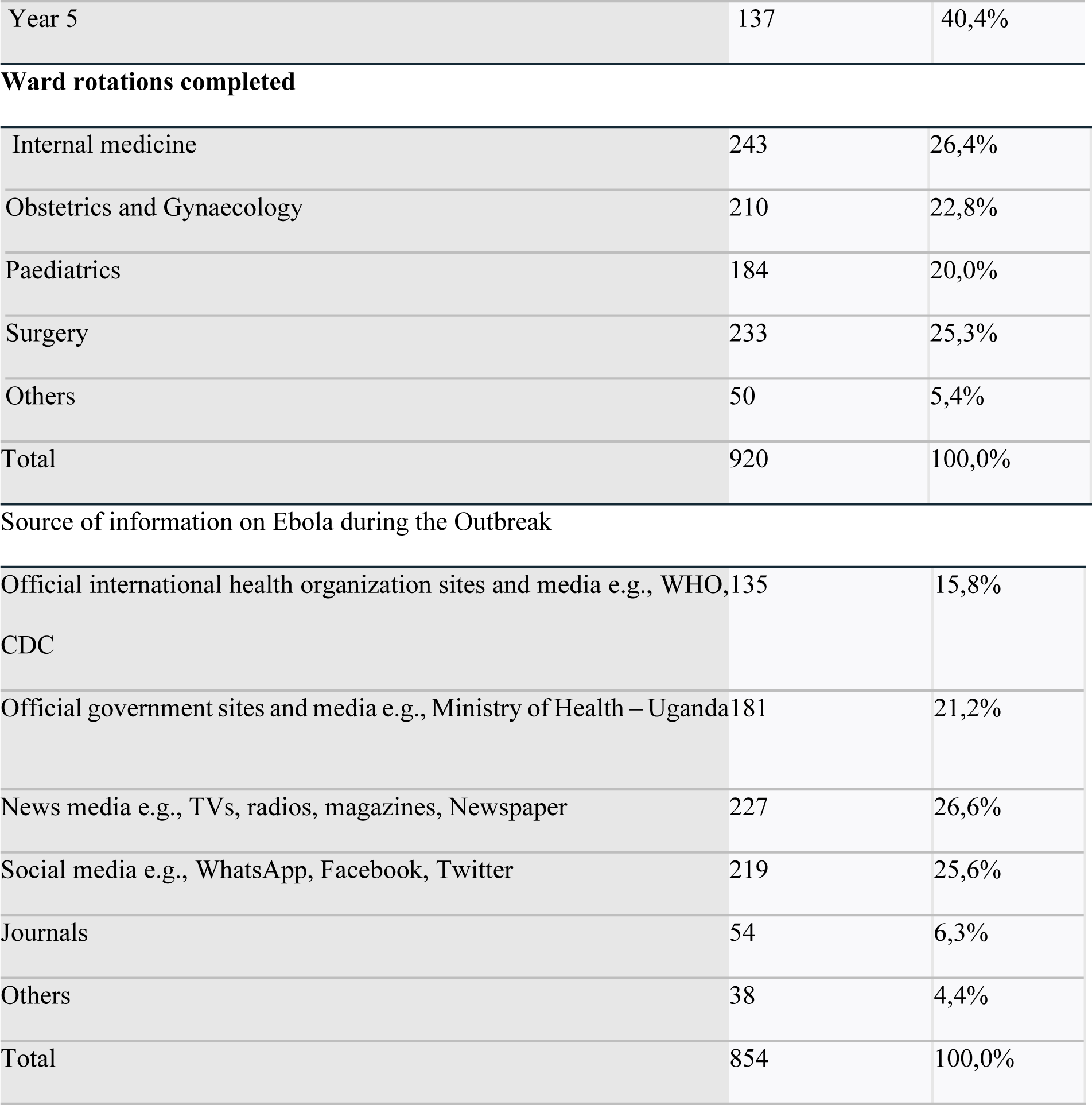
Socio-demographic characteristics of the participants.

| Variable | Freq(n) | Percentage (%) |
| --- | --- | --- |
| **Gender** |  |  |
| Female | 129 | 38,1% |
| Male | 210 | 61,9% |
| **Age (Mean, S.D)** |  |  |
| 24.23, 4.038 |  |  |
| <- 20 | 9 | 2,7% |
| 21-30 | 310 | 91,4% |
| 31-40 | 17 | 5,0% |
| >- 40 | 3 | 0,9% |
| **Course Offered** |  |  |
| Medicine and Surgery (MBChB) | 264 | 77,9% |
| Nursing (BNUR) | 28 | 8,3% |
| Dental Surgery (BDS) | 47 | 13,9% |
| **Year Of Study** |  |  |
| Year 3 | 141 | 41,6% |
| Year 4 | 61 | 18,0% |
| Year 5 | 137 | 40,4% |
| Ward rotations completed |  |  |
| Internal medicine | 243 | 26,4% |
| Obstetrics and Gynaecology | 210 | 22,8% |
| Paediatrics | 184 | 20,0% |
| Surgery | 233 | 25,3% |
| Others | 50 | 5,4% |
| Total | 920 | 100,0% |
| Source of information on Ebola during the Outbreak |  |  |
| Official international health organization sites and media e.g., WHO, CDC | 135 | 15,8% |
| Official government sites and media e.g., Ministry of Health – Uganda | 181 | 21,2% |
| News media e.g., TVs, radios, magazines, Newspaper | 227 | 26,6% |
| Social media e.g., WhatsApp, Facebook, Twitter | 219 | 25,6% |
| Journals | 54 | 6,3% |
| Others | 38 | 4,4% |
| Total | 854 | 100,0% |

### Knowledge

The mean knowledge score was 62.2 (SD: 15.1). 13.6 (n= 46) percent of the participants scored 80% or more were considered to have sufficient knowledge.

The mean knowledge score of female participants was higher than that of male participants (87.2 vs. 83.8%), However this difference was not statistically significant (aOR: 0.8; 95% CI: 0.4–1.9). There was significant association between knowledge and using official international health organisation sites and social media as source of information (aOR: 0.4; 95% CI: 0.2 -0.9). Factors affecting knowledge are highlighted in **Table 2**. 38.1% (n= 129) knew the strain that caused the recent outbreak in Uganda and 50.4% (n= 171) were not sure if there is a vaccine for the strain.

**Table 2.**
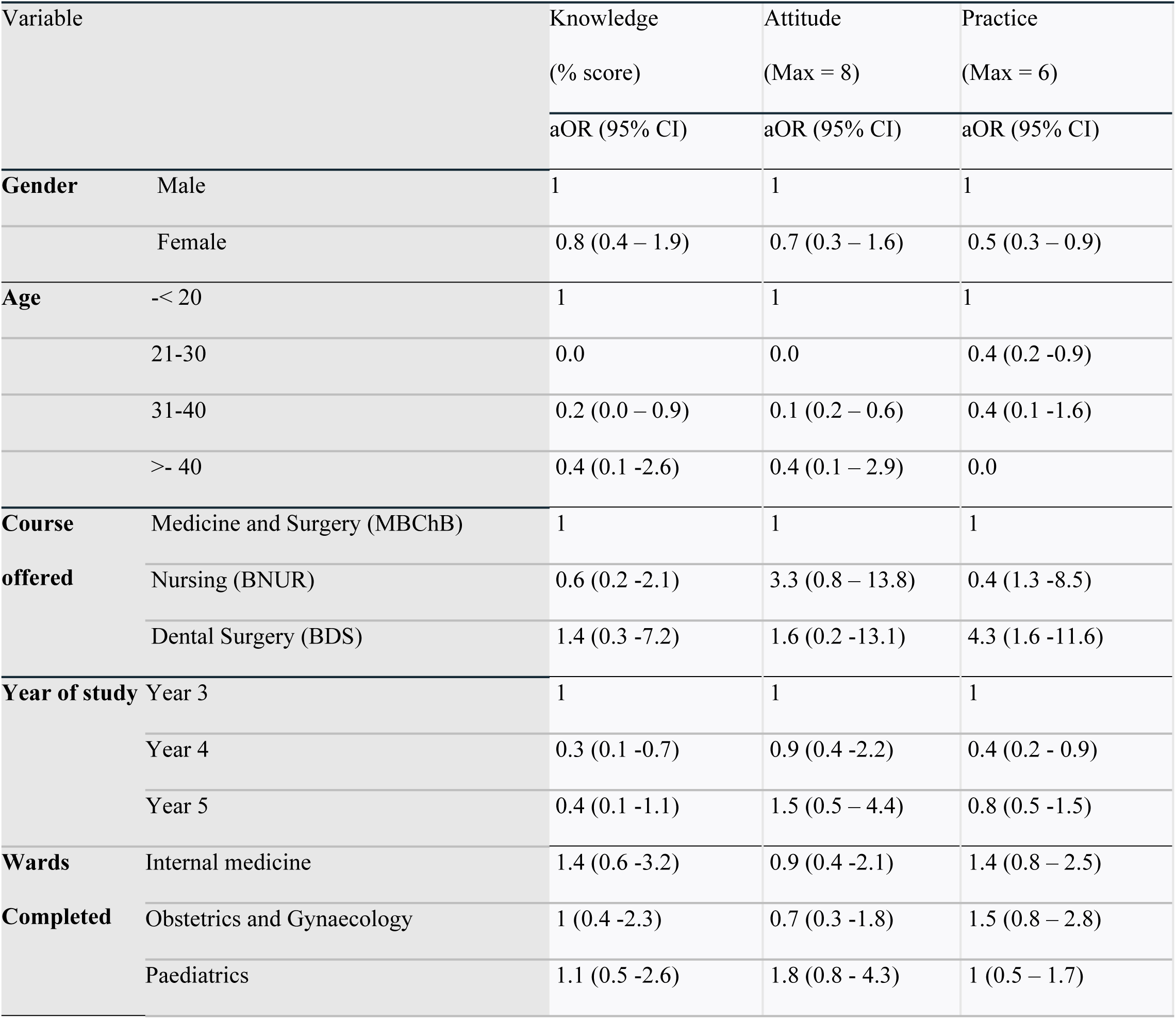
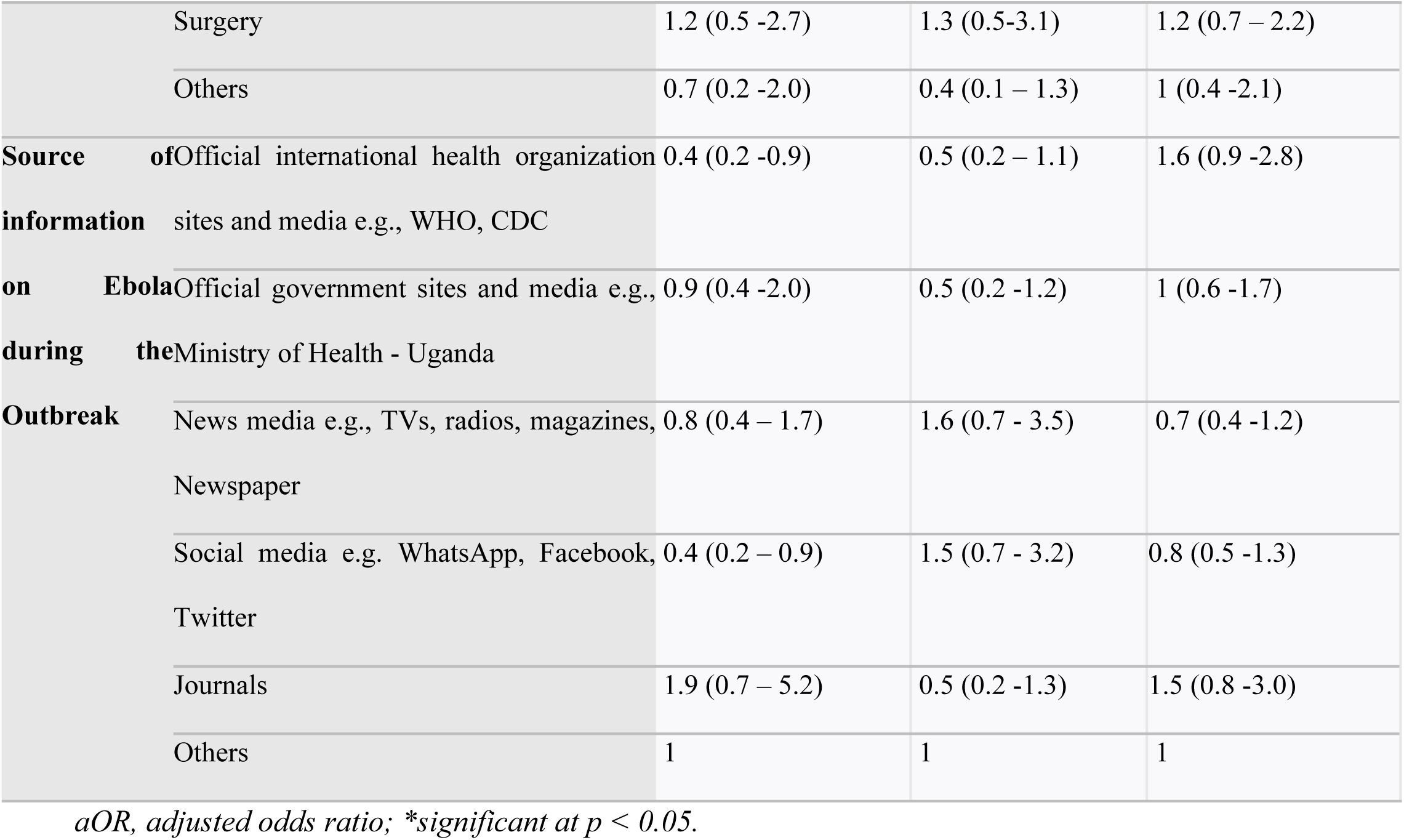
Factors associated with Knowledge, attitude and practices among medical students.

### Attitude

The mean attitude score was 4 (SD: 2.0). Overall, there was poor attitude among participants toward EVD. Only 10.9% (n = 37) of the participants had a good attitude toward EVD. Of these, 73% (n= 27), 86.5% (n= 32) and 45.9% (n= 17) were male participants, pursuing MBChB and in year 5 respectively. 11.8% (n= 40) would confidently participate in the management of a patient with EVD and 41.6% (n= 141) would accept an approved Ebola vaccine for themselves. When asked about the need for training, 55.2% ( n= 187) reported that government and other institutions should include medical students in special training to handle outbreaks in the country. Additionally, 39.5% (n= 134) believe that medical students in teaching hospitals are part of frontline healthcare workers. **Table 3** shows the frequency of the responses and **Table 4** shows the mean attitude scores and percentage of participants with good attitude.

**Table 3.** Attitudes of medical students towards EVD.

| Please indicate the extent to which you agree with the following statement: | Strongly Disagree<br>(n, %) | Disagree<br>(n, %) | Not Sure<br>(n, %) | Agree<br>(n, %) | Strongly Agree<br>(n, %) |
| --- | --- | --- | --- | --- | --- |
| Ebola is a serious Disease* | 1(0.3%) | 2(0.6%) | 1(0.3%) | 10 (2.9%) | 325 (95.9%) |
| Ebola can be prevented* | 39 (11.5) | 14(4.1) | 11 (3.2) | 69 (20.4) | 206 (60.8) |
| I was scared of getting Ebola* | 45 (13.3) | 21 (6.2) | 15 (4.4) | 69 (20.4) | 189 (55.8) |
| Medical Students are part of the frontline workers* | 56 (16.5) | 37 (10.9) | 41 (12.1) | 71 (20.9) | 134 (39.5) |
| When a patient has signs and symptoms of Ebola, I can confidently participate in the management of the patient. * | 85 (25.1) | 72 (21.2) | 82 (24.2) | 60 (17.7) | 40 (11.8) |
| If there was an approved vaccine that could prevent Ebola, I would accept it for myself. * | 37 (10.9) | 32 (9.4) | 53 (15.6) | 76 (22.4) | 141 (41.6) |
| I am familiar with personal protective equipment, donning and doffing procedure* | 44 (13.0) | 38 (11.2) | 44 (13.0) | 75 (22.1) | 138 (40.7) |
| The government and other institutions should include medical students in special trainings to handle outbreaks in the country* | 42 (12.4) | 22 (6.5) | 27 (8.0) | 61 (18.0) | 187 (55.2) |

**Table 4.**
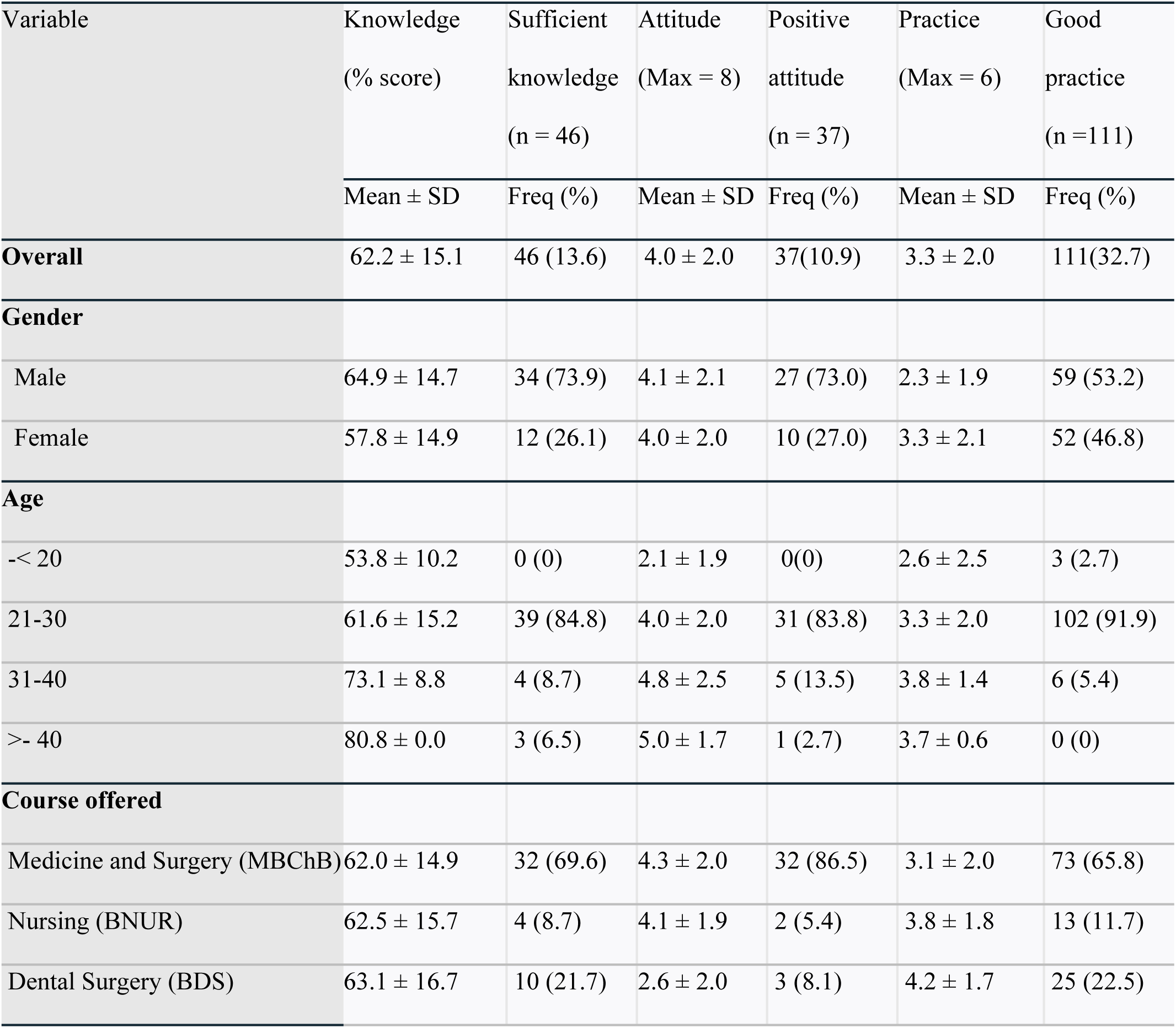
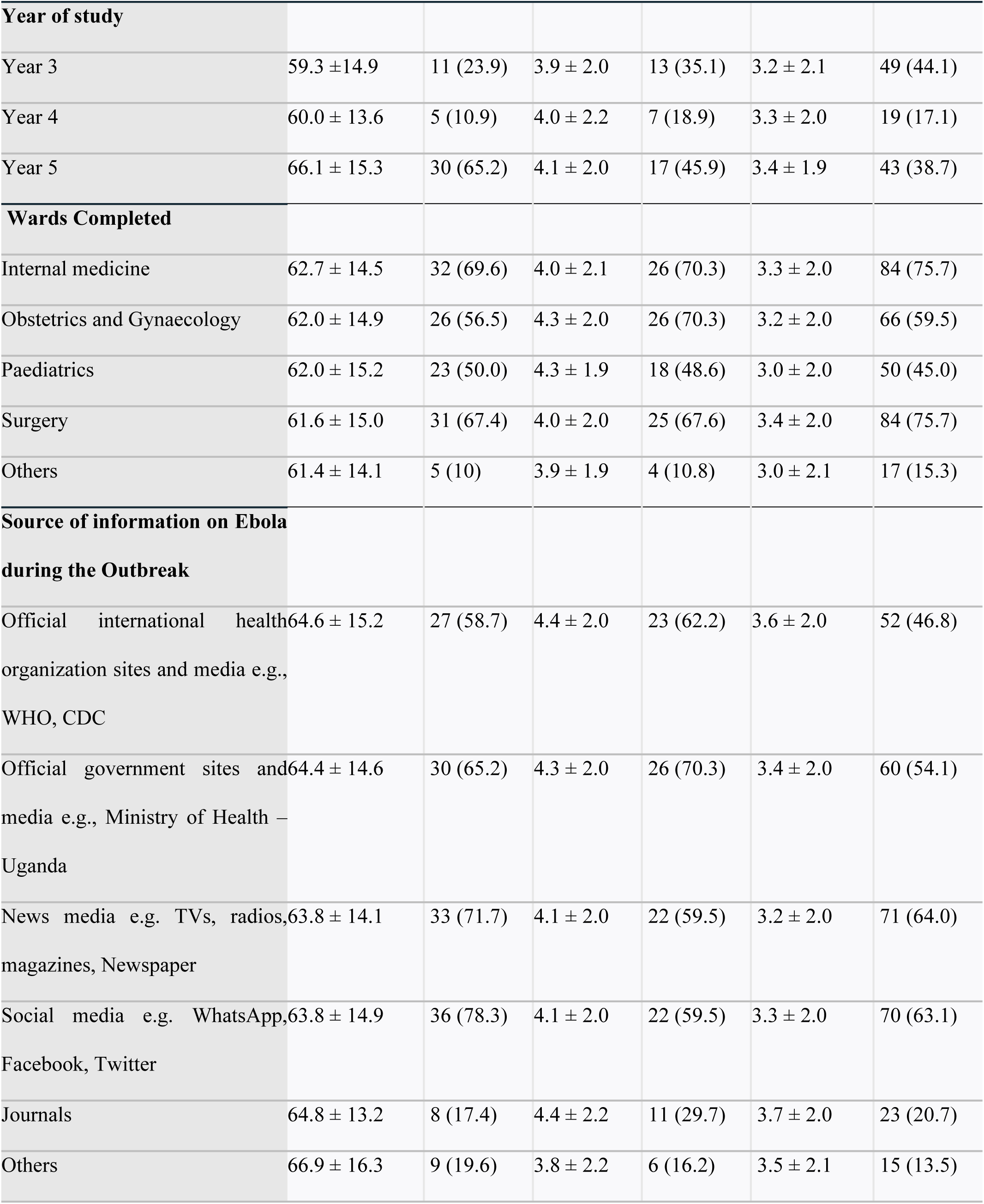
Knowledge, attitude and practices among medical students.

| Variable | Knowledge | Sufficient | Attitude | Positive | Practice | Good |
| --- | --- | --- | --- | --- | --- | --- |
|  | (% score) | knowledge | (Max = 8) | attitude | (Max = 6) | practice |
|  |  | (n = 46) |  | (n = 37) |  | (n = 111) |
| | Mean ± SD | Freq (%) | Mean ± SD | Freq (%) | Mean ± SD | Freq (%) |
| **Overall** | 62.2 ± 15.1 | 46 (13.6) | 4.0 ± 2.0 | 37(10.9) | 3.3 ± 2.0 | 111(32.7) |
| **Gender** |  |  |  |  |  |  |
| Male | 64.9 ± 14.7 | 34 (73.9) | 4.1 ± 2.1 | 27 (73.0) | 2.3 ± 1.9 | 59 (53.2) |
| Female | 57.8 ± 14.9 | 12 (26.1) | 4.0 ± 2.0 | 10 (27.0) | 3.3 ± 2.1 | 52 (46.8) |
| **Age** |  |  |  |  |  |  |
| < 20 | 53.8 ± 10.2 | 0 (0) | 2.1 ± 1.9 | 0(0) | 2.6 ± 2.5 | 3 (2.7) |
| 21-30 | 61.6 ± 15.2 | 39 (84.8) | 4.0 ± 2.0 | 31 (83.8) | 3.3 ± 2.0 | 102 (91.9) |
| 31-40 | 73.1 ± 8.8 | 4 (8.7) | 4.8 ± 2.5 | 5 (13.5) | 3.8 ± 1.4 | 6 (5.4) |
| > 40 | 80.8 ± 0.0 | 3 (6.5) | 5.0 ± 1.7 | 1 (2.7) | 3.7 ± 0.6 | 0 (0) |
| **Course offered** |  |  |  |  |  |  |
| Medicine and Surgery (MBChB) | 62.0 ± 14.9 | 32 (69.6) | 4.3 ± 2.0 | 32 (86.5) | 3.1 ± 2.0 | 73 (65.8) |
| Nursing (BNUR) | 62.5 ± 15.7 | 4 (8.7) | 4.1 ± 1.9 | 2 (5.4) | 3.8 ± 1.8 | 13 (11.7) |
| Dental Surgery (BDS) | 63.1 ± 16.7 | 10 (21.7) | 2.6 ± 2.0 | 3 (8.1) | 4.2 ± 1.7 | 25 (22.5) |

| **Year of study** |  |  |  |  |  |  |
| --- | --- | --- | --- | --- | --- | --- |
| Year 3 | 59.3 ± 14.9 | 11 (23.9) | 3.9 ± 2.0 | 13 (35.1) | 3.2 ± 2.1 | 49 (44.1) |
| Year 4 | 60.0 ± 13.6 | 5 (10.9) | 4.0 ± 2.2 | 7 (18.9) | 3.3 ± 2.0 | 19 (17.1) |
| Year 5 | 66.1 ± 15.3 | 30 (65.2) | 4.1 ± 2.0 | 17 (45.9) | 3.4 ± 1.9 | 43 (38.7) |
| **Wards Completed** |  |  |  |  |  |  |
| Internal medicine | 62.7 ± 14.5 | 32 (69.6) | 4.0 ± 2.1 | 26 (70.3) | 3.3 ± 2.0 | 84 (75.7) |
| Obstetrics and Gynaecology | 62.0 ± 14.9 | 26 (56.5) | 4.3 ± 2.0 | 26 (70.3) | 3.2 ± 2.0 | 66 (59.5) |
| Paediatrics | 62.0 ± 15.2 | 23 (50.0) | 4.3 ± 1.9 | 18 (48.6) | 3.0 ± 2.0 | 50 (45.0) |
| Surgery | 61.6 ± 15.0 | 31 (67.4) | 4.0 ± 2.0 | 25 (67.6) | 3.4 ± 2.0 | 84 (75.7) |
| Others | 61.4 ± 14.1 | 5 (10) | 3.9 ± 1.9 | 4 (10.8) | 3.0 ± 2.1 | 17 (15.3) |
| **Source of information on Ebola during the Outbreak** |  |  |  |  |  |  |
| Official international health organization sites and media e.g., WHO, CDC | 64.6 ± 15.2 | 27 (58.7) | 4.4 ± 2.0 | 23 (62.2) | 3.6 ± 2.0 | 52 (46.8) |
| Official government sites and media e.g., Ministry of Health – Uganda | 64.4 ± 14.6 | 30 (65.2) | 4.3 ± 2.0 | 26 (70.3) | 3.4 ± 2.0 | 60 (54.1) |
| News media e.g. TVs, radios, magazines, Newspaper | 63.8 ± 14.1 | 33 (71.7) | 4.1 ± 2.0 | 22 (59.5) | 3.2 ± 2.0 | 71 (64.0) |
| Social media e.g. WhatsApp, Facebook, Twitter | 63.8 ± 14.9 | 36 (78.3) | 4.1 ± 2.0 | 22 (59.5) | 3.3 ± 2.0 | 70 (63.1) |
| Journals | 64.8 ± 13.2 | 8 (17.4) | 4.4 ± 2.2 | 11 (29.7) | 3.7 ± 2.0 | 23 (20.7) |
| Others | 66.9 ± 16.3 | 9 (19.6) | 3.8 ± 2.2 | 6 (16.2) | 3.5 ± 2.1 | 15 (13.5) |

### Practices

Overall, 32.7% (n= 111) had good practices with mean score 3.3 (SD: 2.0). Among the participants, 64.3% (n= 218) wore gloves while attending to patients and 68.1% (n= 231) washed hands before and after handling patients. Unfortunately, 54.9% (n= 186) avoided patients with signs and symptoms suggestive of EVD. **Table 5** shows practices of medical students.

**Table 5.** Preventive Practices of medical students.

| How often did you do the Following? | Always (n, %) | Occasional (n, %) | Never (n, %) |
| --- | --- | --- | --- |
| In the recent Ebola outbreak, I was avoiding crowded places. * | 168 (49.6) | 130 (38.3) | 41 (12.1) |
| In the recent Ebola outbreak, I was wearing a mask or face shield when in contact with patients. * | 145 (42.8) | 117 (34.5) | 77 (22.7) |
| In the recent Ebola outbreak, I was wearing gloves when in contact with patients* | 218 (64.3) | 89 (26.3) | 32 (9.4) |
| In the recent Ebola outbreak, I was washing hands before and after handling patients* | 231 (68.1) | 78 (23.0) | 30 (8.8) |
| In the recent Ebola outbreak, I was refraining from shaking hands* | 166 (49.0) | 126 (37.2) | 47 (13.9) |
| In the recent Ebola outbreak, I avoided patients with signs and symptoms suggestive of Ebola* | 186 (54.9) | 97 (28.6) | 56 (16.5) |

## Discussion

The mean knowledge score of 62.2% on questions about knowledge indicating moderate knowledge among the participants. This was higher than a study done among students at a school of medicine in Dakar, Senegal (39.6%) (14) and a score of 54.7% among nursing students in Yangon (15). However, this score in much lower than among students of a college in Education in Lagos, Nigeria (94%)(16).

Our study showed that information about EVD was mainly obtained through News media (e.g. TVs, radio, magazines and newspaper), social media ( e.g. WhatsApp, twitter and Facebook) and official government sites and media like Ministry of health. This was comparable to a study in among nursing students where information about EVD was obtained through TV, radio, internet news and peers (15).

Bleeding (92.3%), fever (81.7%), vomiting (79.9%), diarrhea (75.2%), weakness (71.7%) and headache (65.2%) were the most recognised signs of EVD. This is similar to a study on medical students in Dakar where Fever (86.3%), diarrhea (77.1%), vomiting (73.9%), headache (64.3%) and abnormal bleeding (62.6%) were the main features reported by students (14). The diversity and non-specific signs and symptoms of EVD can mimic many tropical diseases despite the good knowledge about clinical signs from our study.

Physical contact (75.2%), shaking hands (67.3%) and secretions from an infected person i.e. blood (88.5%), sweat (81.7%), urine (63.4%) and sperms and vaginal fluid (65.8%) were the most identified mode of transmission of EVD. A study in Pakistan among medical students of Rawalpindi, found that students were aware that EVD can be transmitted via body fluids; saliva (52.6%), blood (55.8%), tears (39.6%), semen (41.6%) and dead body (47.4%) (17). This knowledge is vital in reducing the risk of transmission of EVD. 80.2% of our participants believe that one can survive and recover from EVD. This can compared to 73% from Mbaye’s study (14) and 68% reported by Mbuk (18). Less than half of the participants (38.1%) knew that Sudan ebolavirus was responsible form the recent outbreak and 50.4% were not sure if there was a vaccine for this strain of ebolavirus.

### Attitude

In our study, majority of the participants (95.9%) reported EVD to be a serious disease and only 11.8% would participate in management of a patient with the disease. This was in congruent with study in Sierra Leon where 95% showed discriminatory attitudes towards EVD survivors (19). Our study also indicated that 41.6% would take an approved ebolavirus vaccine while 40.7% were familiar with personal protective equipment, donning and doffing procedures. Additionally, more than half (55.2%), believe that there is need for training of medical students on how to handle outbreaks like EVD. This will improve not only their attitudes but also knowledge and practices towards disease prevention and management.

### Practices

This study indicated that washing hands regularly (68.1%) and using gloves while handling patients (64.3%) were the mostly applied measures against EVD during the outbreak. Others reported avoiding crowded places (49.6%), avoiding handshake (49%) and wearing face mask or shield when in contact with patients (42.8%). However, 54.9% reported avoiding patients with signs and symptoms suggestive of EVD. The inadequacy of personal protective equipment could explain why they desisted from attending to such patients.

## Conclusion

These findings highlight concerns regarding knowledge, attitude, and practice among medical students regarding EVD outbreak. Therefore, our findings will help policy and decision making institutions to plan and develop new interventions to improve students’ knowledge, attitude, and practice towards future outbreaks.

## Ethical consideration

The proposal was approved by The Aids Support Organisation Research and Ethics Committee (TASO REC) Protocol Number TASO-2023-250. All participants signed a written informed consent.

## Data Availability

All relevant data are within the manuscript and its supporting information files.

## Acknowledgement

We would like to acknowledge the following people for their significant support and contribution towards this publication; Health Professions Education and Training for Strengthening the Health Systems and Services in Uganda (HEPI-SHUSS) for training on proposal writing and publication, Eddy Kyagulanyi for proposal development, Jaimin Varsani for data analysis, Beliza Chemutai for data collection, Elizabeth Kiyingi Nakiyingi and Derrick Bary Abila.

